# Atmosome: The Personal Atmospheric Exposome

**DOI:** 10.1101/2020.07.02.20145433

**Authors:** Hari Bhimaraju, Vaibhav Pandey, Nitish Nag, Ramesh Jain

## Abstract

Modern health research is increasingly separated into various “omics” fields. Omics provides a quantified approach to the total collective information about the various realms of biology. One aspect of omics called the exposome relates to the collective exposures an individual is placed in throughout their life. A subset of this exposome, is the atmospheric exposome, which we name as the “atmosome”. Air surrounds individuals from the moment they are born till they take their last breath. The atmosphere is constantly changing, given our location and surroundings are always dynamic throughout life. The atmosphere inevitably plays a massive role in our health such as impact on DNA damage, metabolism, skin integrity, and lung health. In this work, we discuss the significance of the atmosome in personalized health and present our atmosome sensing system. We developed and analyzed data from our IoT, microcontroller-based system which collects real-time individual air quality data and posts it to a cloud server for immediate access. Our experimental results demonstrate the accuracy of the data we collected and the avenues our system creates for direct lifestyle to environment correlations. Quantifying the individual atmosome is a next step in advancing personalized health and medical research. Our two main goals in this paper are to present the atmosome as a concept and to explain how to track it using low-cost electronics.

## I. Introduction

“Our most basic common link is that we all inhabit this planet. We all breathe the same air. We all cherish our children’s future. And we are all mortal.”

- John F. Kennedy

Health is a constantly changing state that is affected by various internal and external factors, including the genome, microbiome, and exposome. An individual’s exposome consists of all of their exposures, from their time in their mother’s womb through the rest of their life. It considers lifestyle factors, occupational and socioeconomic conditions, and environmental settings to develop an in-depth understanding of how an individual’s surroundings directly impact their health state. The atmospheric exposome focuses on the effects of the environment, and specifically air, on health. Air surrounds everyone, no matter where they live or what they are doing. Thus, it inevitably plays a crucial role in determining health. Americans spend approximately 90 percent of their time indoors, where the concentrations of some air pollutants are 2 to 5 times higher than in the outdoors [8]. This poor indoor air quality can cause various infections, lung cancer, and chronic lung diseases such as asthma [1]. It also can contribute to the development of atherosclerosis, an underlying cause for many cardiovascular diseases [30]. The detrimental health effects of air quality are just as prominent in the outdoors. Ambient air pollution was responsible for 3 million deaths worldwide in 2017, and it is the single biggest environmental health risk [32]. Annually, 9 out of every 10 people breathe air containing high levels of pollutants [24]. To improve air quality and minimize pollution-related deaths, we must identify, measure, and analyze our atmosome to study how the air we breathe affects our health. Exposome research is expanding rapidly and public environmental data is becoming more detailed and accessible. However, the need for a personal, real-time, multi-stream air measurement system from which researchers can easily mine data is still unsolved.

Currently, there are no research investigations that characterize the body’s interactions with air as an “omics” field. Thus, we coined the word atmosome to describe the atmospheric sub-component of an individual’s exposome. Broadly, the motivation for this work is to use the atmosome to further personalized health state estimation from multi-modal data [18], [15]. Continued research in this field will look into expanding to other health domains, improving quality metrics, tackling performance issues, and developing methods to combine the atmosome data with other data streams. Finding unique relationships between these data streams allows for better models of the user’s individual lifestyle. In the atmosome case, this allow us to find the impact of different atmospheric and environmental data streams on behavioral outcomes such as sleep using appropriate N=1 modelling techniques [28].

## II. Related Work

Before assessing the novelty and efficacy of the atmosome data collection system, we must become familiar with existing air quality measurement systems. In this section, we will survey current solutions in the market and discuss their uses and shortcomings.

The EPA gathers outdoor air pollution data from several federal, state, and local agencies and stores it in a database called the Air Quality System (AQS). However, these monitoring systems are usually only installed in urban areas and take measurements for only some air pollutants every few days. Moreover, they do not consider indoor air quality or assess the living conditions of specific individuals [7]. Thus, the data in the AQS is pretty limited.

Some existing commercial air monitoring systems include Dyson Pure Hot+Cool(tm) HP04 purifying heater + fan tool, Molekule, Honeywell True HEPA Air Filter, and Alen BreatheSmart 75i [11] [10]. The Dyson, which costs $649.99, uses HEPA filters and senses airborne particles (PM 2.5 and 10) and gases (VOC and NO2). It comes with a smart app feature. However, it lacks the capability to destroy Formaldehyde, is unable to sense very small particles (i.e. 0.1 microns), is not approved by the Association of Home Appliance Manufacturers, and is very expensive [6]. Molekule, priced at $799, actively filters the air using Photo Electrochemical Oxidation (PECO) filters to destroy pollutants at the molecular level. It kills RNA/DNA viruses, VOCs, and more, but its price is still quite high for many potential buyers. It also has no air temperature controls, is not AHAM (Association of Home Appliance Manufacturers) approved, and is not HEPA filter-equipped [13].

As with the aforementioned examples, listed in Table I, most commercial air monitoring systems tend to be very expensive and are not geared towards research. They lack a cloud component to easily post and retrieve time-stamped data and focus on air purification as opposed to varied air stream data collection. Most significantly, they are not accessible to most individuals due to their prices. AMS is built with these characteristics and costs between $83 and $107, making it significantly cheaper than the industry alternatives.

**Table I.** Examples of existing air quality measurement and cleansing products in the industry.

| Product | Price | Measurements |
| --- | --- | --- |
| Dyson Pure Hot+Cool™ | \$649.99 | PM2.5, PM10, VOC, NO2, Formaldehyde |
| Molekule | \$799 | RNA/DNA viruses, VOCs, Formaldehyde, allergens, and more |
| Honeywell | \$179.99 | Odors, VOCs, certain germs |
| Alen BreatheSmart | \$749 | Particles larger than 0.3 microns |

## III. Personal Atmosome Sensor System

AMS (Atmosome Measurement System) is a low-cost tool for capturing raw data on individuals’ atmospheric exposomes. This IoT air monitoring kit considers geospatial data to track air quality from seventeen different streams. It enables the user’s environment to be monitored over an indefinite period and posts their data to the cloud in real-time. These features allow for easy atmosome monitoring without interfering with a user’s natural lifestyle, which makes it simple for users or researchers to connect their actions at every moment to changes in their environment. They can then use this data to generate reports, study action-to-environment patterns, and make algorithms to predict behavioral or environmental triggers for atmosome state. Another advantage of the system being cloud-based is that it allows researchers to compare and study data from different users around the world, or even from the same user during travel, to analyze correlations between lifestyle and atmosome.

AMS comes in two variations: a portable version with a reduced number of sensors that charges via a USB cable and laptop or a complex version that plugs into a wall outlet for indoor room measurements. The former version costs approximately $84 and the latter version costs approximately $108. Figure 2 shows the advanced model.

**Fig 1.**
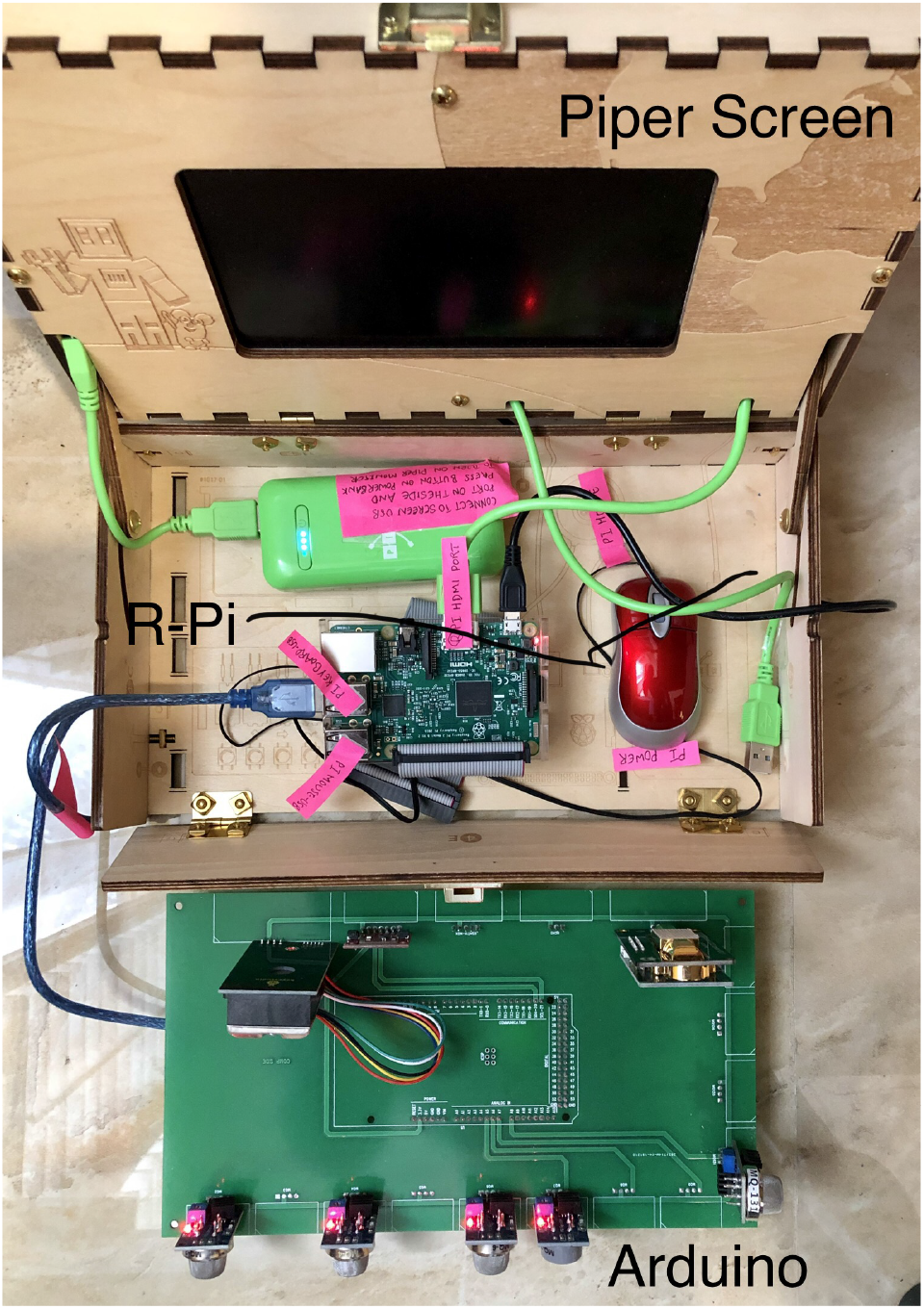
System hardware, including a monitor in a Piper Kit box, a Raspberry Pi, an Arduino, and a PCB.

**Fig 2.**
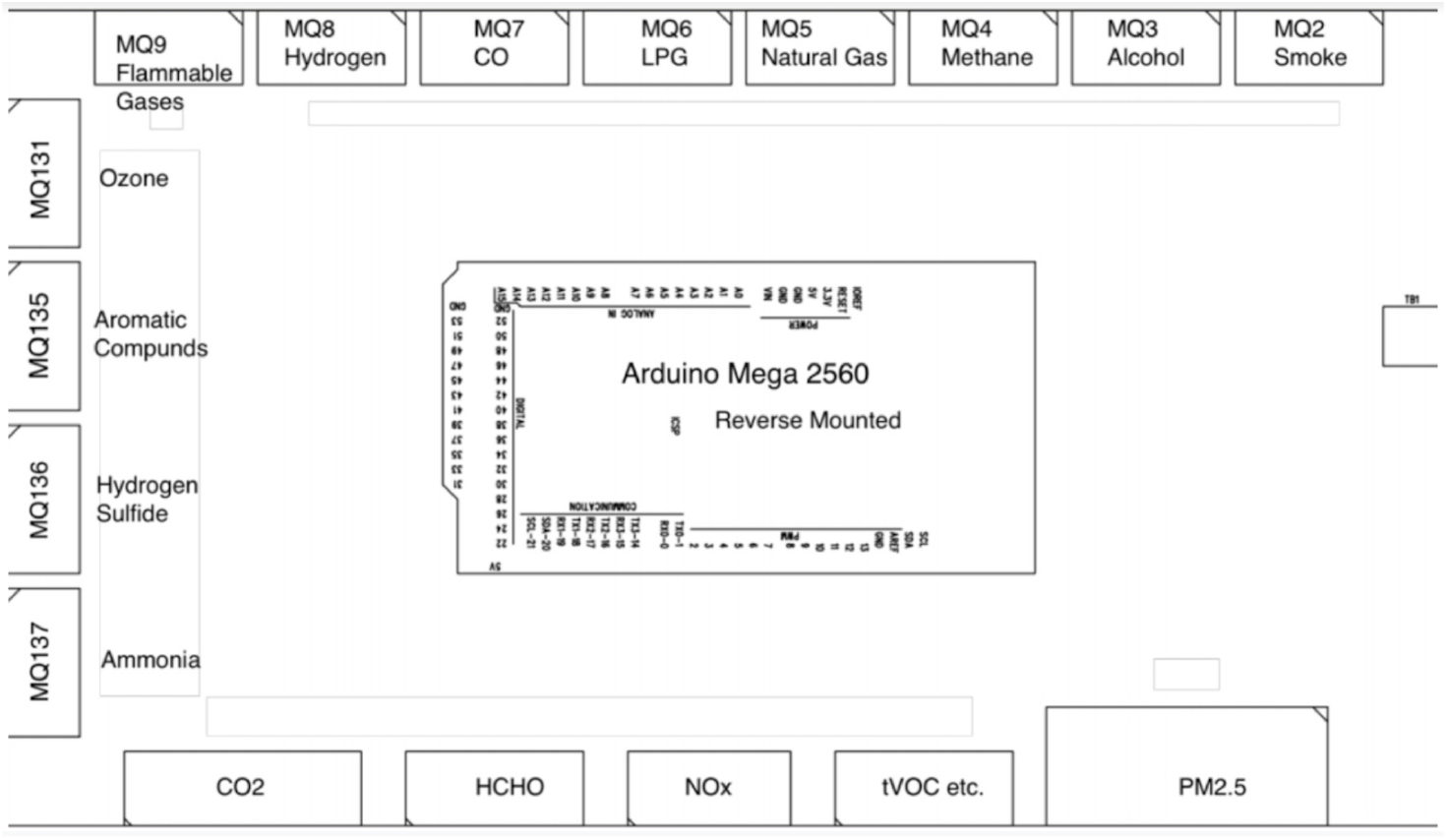
Circuit diagram of the PCB:, This board holds the Arduino and 17 environmental sensors.

The Arduino Mega powers the sensors, collects sensor data, and sends the readings to a Raspberry Pi. The Pi records this data onto the cloud. The cloud system is built using a Python Flask framework and provides a REST API for retrieving and posting data. It also comes with an API to query based on specific locations. Users and researchers can easily collect and parse this data to generate graphs and study patterns over time. Figure 3 shows the two aspects of the system architecture: hardware components and cloud mechanisms. Figure 1 displays a photograph of AMS.

**Fig 3.**
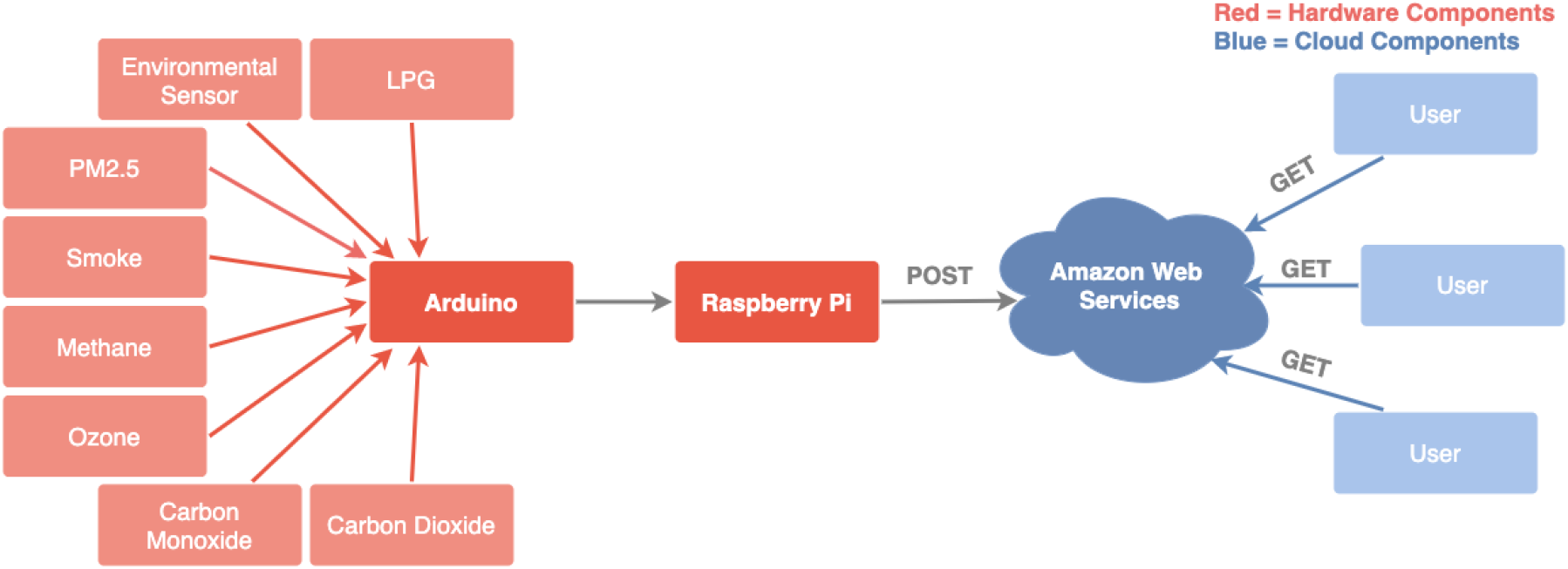
System architecture chart: Red denotes hardware components while blue represents cloud computing components

The air quality system includes sensors and microcontrollers which measure various air streams, listed in Table II.

**Table II.** Information about hardware components of AMS, including sensors and microcontrollers

| Sensor | Measurements | Producer | Type |
| --- | --- | --- | --- |
| CCS811/BME280 | tVOC, eCO<sub>2</sub>, altitude, temp, pressure | Sparkfun | Environmental combo sensor |
| MQ2 | Smoke | Keywish | Metal oxide Semiconductor (MOS) gas sensor |
| MQ3 | Alcohol | Keywish | MOS gas sensor |
| MQ4 | Methane | Keywish | MOS gas sensor |
| MQ5 | Natural gas | Keywish | MOS gas sensor |
| MQ6 | Liquefied Petroleum Gas | Keywish | MOS gas sensor |
| MQ7 | Carbon Monoxide | Keywish | MOS gas sensor |
| MQ8 | Hydrogen | Keywish | MOS gas sensor |
| MQ9 | Flammable Gases | Keywish | MOS gas sensor |
| MQ131 | Ozone | SainSmart | MOS gas sensor |
| MQ135 | Aromatic Compounds, Sulfide | SainSmart | MOS gas sensor |
| MQ136 | Hydrogen Sulfide | SainSmart | MOS gas sensor |
| MQ137 | Ammonia | SainSmart | MOS gas sensor |
| NOx | Nitrogen Oxides | Ogam | MOS gas sensor |
| HCHO | Formaldehyde | Gravity | MOS gas sensor |
| CO<sub>2</sub> | Carbon Dioxide | DFRobot | NDIR |
| PM2.5 | Particulate Matter | Keyestudio | Optical, IR emitting diode |
| Arduino Mega 2560 | Reads all the sensors values periodically and performs calculations | Arduino | Microcontroller |
| Raspberry Pi | Receives data from Arduino using a Python program and calls REST API to the cloud | Raspberry Pi | Microcontroller |

**Table III.** Environmental parameters and reference ranges.

| Parameter | Good | Average | Poor | Bad |
| --- | --- | --- | --- | --- |
| Humidity | 30–50% | 50–60% | <30%, >60% | <20%, >70% |
| Temperature | 20–24°C (winter) 24–27°C (summer) | 16–20°C, 27–28°C | 28–30°C | <16°C, >30°C |
| PM2.5 | 0–12 $\mu\text{g}/\text{m}^3$ | 12–15 $\mu\text{g}/\text{m}^3$ | 15–35 $\mu\text{g}/\text{m}^3$ | >35 $\mu\text{g}/\text{m}^3$ |
| tVOC | 0–220 ppb | 221–660 ppb | 661–2000 ppb | >2000 ppb |
| O<sub>3</sub> | 0.05 ppm | 0.08 ppm | 0.1 ppm | >= 0.2 ppm |
| CO<sub>2</sub> | 350–1000 ppm | 1001–2000 ppm | 2001–5000 ppm | >5000 ppm |
| CO | 0–3 ppm | 3–8 ppm | 8–10 ppm | >10 ppm |

AMS collects data from seventeen sensor streams. Each gas has unique effects on health and different threshold values for what is considered dangerous for humans. Understanding the basics of the major factors the system measures is crucial to utilizing its data for lifestyle and health state analysis.

## IV. Experimental Results

We conducted various experiments to test the sensor kit in both outdoor and indoor conditions. In this paper, we discuss the parameters displayed in III discussion. Green lines represent optimal healthy values, red lines represent dangerous or unhealthy values, and yellow lines represent sub-optimal values.

### A. Temperature and Humidity

Figures 4 and 5 show graphs of humidity and temperature measurements taken from three different locations, including a household and minimal travel in the US, a household and minimal travel in India, and an airplane flight of 17 hours. The low humidity readings from the plane confirm dryness and discomfort effects felt in air travel and align with the United States CDC’s air travel yellow book [7].

**Fig 4.**
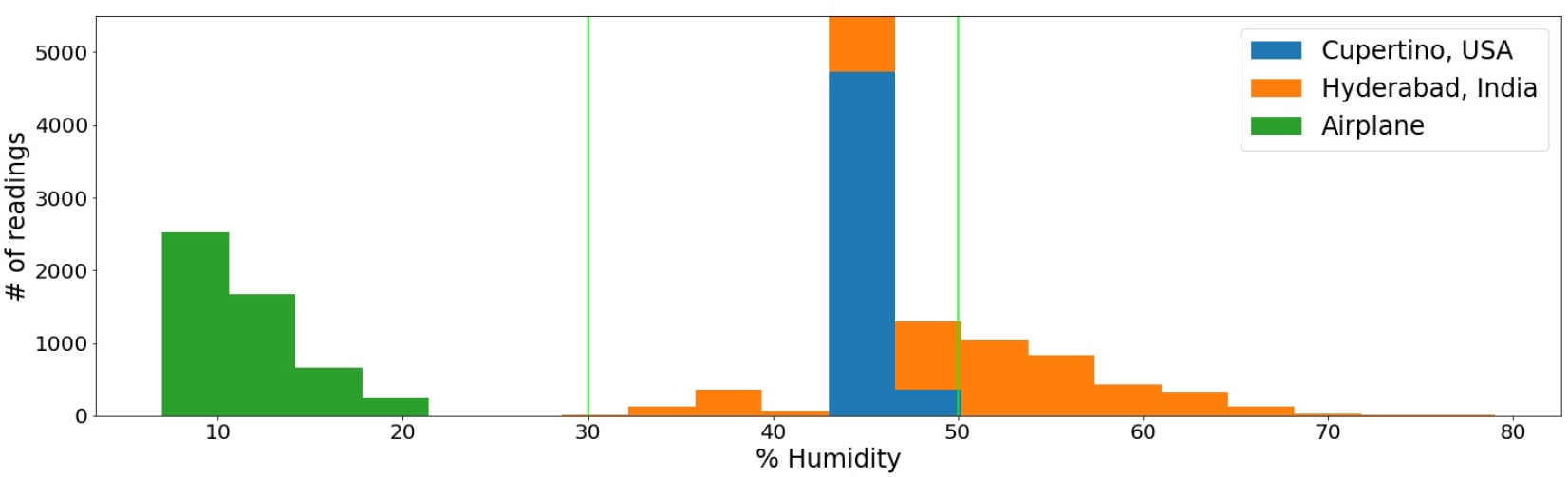
Humidity (%): These are color coded humidity conditions of the user atmospheric home conditions in Cupertino, California, Hyderabad, India, and a commercial airplane. 30% to 50% marks the healthy range (green vertical lines). We can clearly see the effect on the atmosome humidity during air travel.

**Fig 5.**
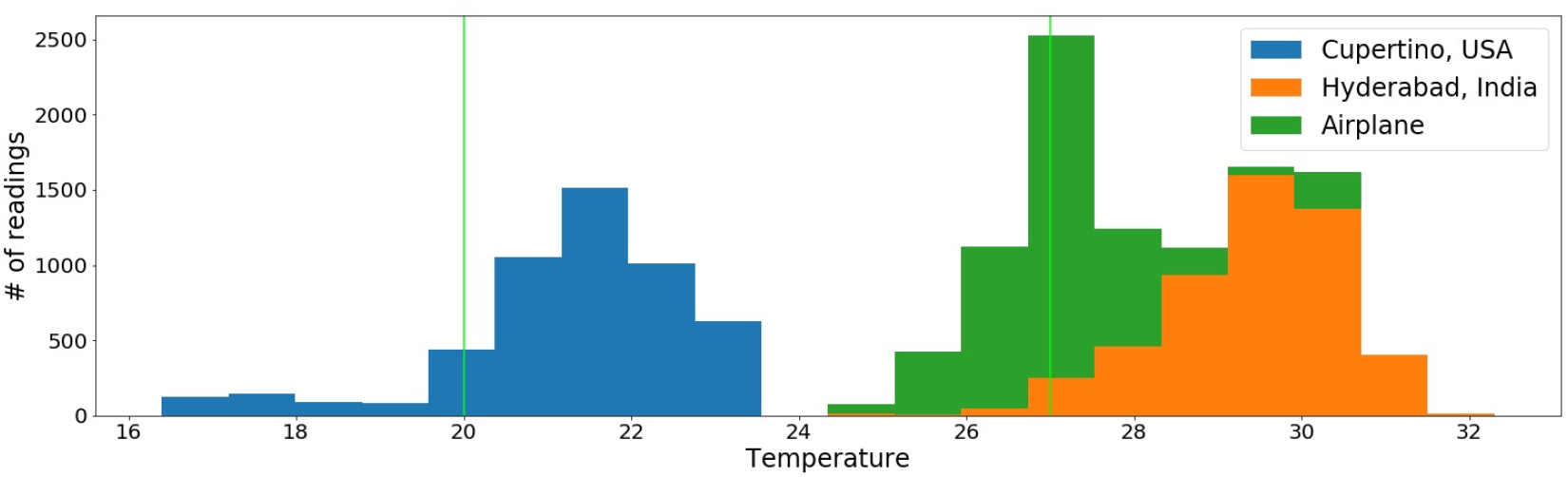
Temperature (Celsius): User atmospheric temperature at home in Cupertino, California, Hyderabad, India, and a commercial airplane. 20 degrees to 27 degrees marks a comfortable and healthy range (green vertical lines).

A second sensor kit in another user location gathered data yielding the time series data shown in Figures 6 and 7.

**Fig 6.**
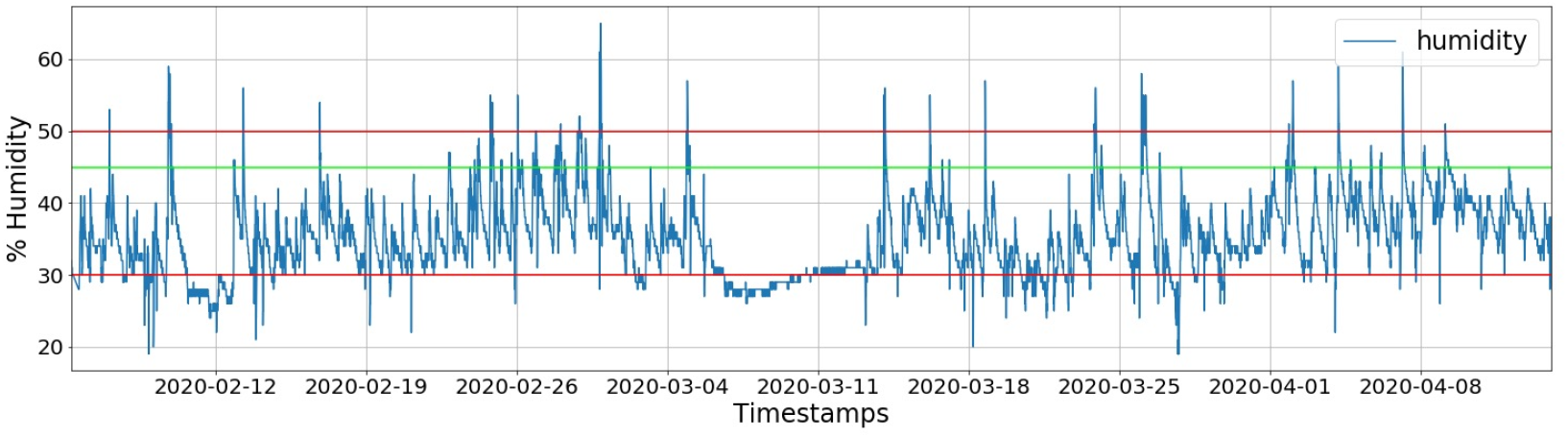
Humidity (%): User home from February to April 2020 at home in the Sierra Mountains, El Dorado County, California. 45% is the optimal value (green), while 30% (red) to 50% (pink) is the healthy range.

**Fig 7.**
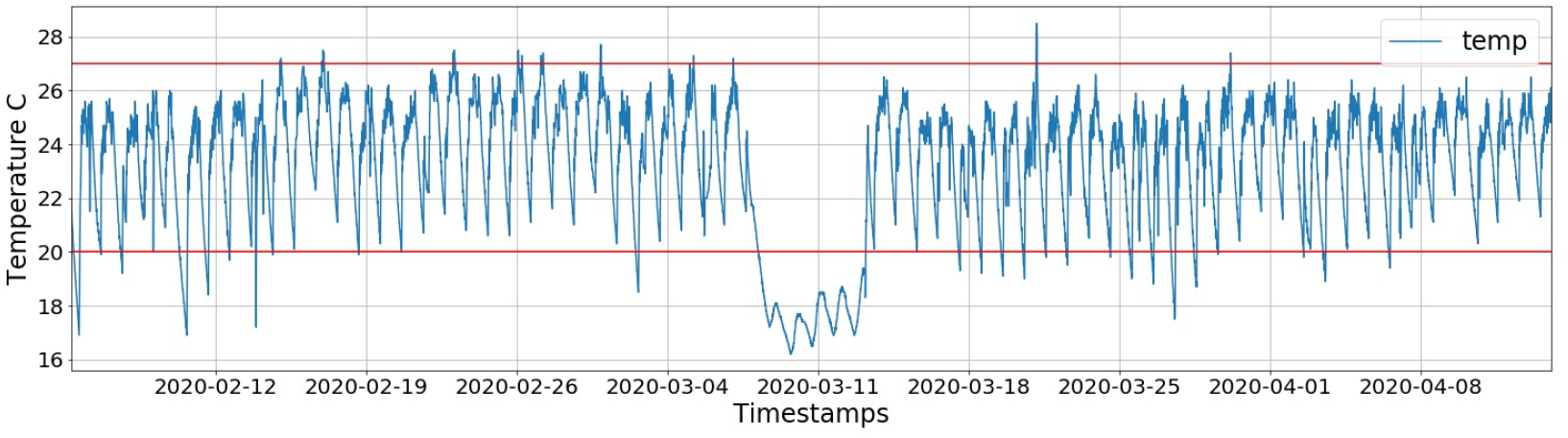
Temperature (Celsius): User home from February to April 2020 at home in the Sierra Mountains, El Dorado County, California. 20 degrees to 27 degrees marks the healthy range (red horizontal lines).

### B. Carbon Dioxide

Figure 8 shows a graph of the CO_2_ measurements. The results were different than expected because the readings in the US showed much higher indoor CO_2_ levels than the readings in a more polluted area in India. Further analysis revealed, however, that the closed windows and doors throughout the day in wintery US weather reduced circulation and increased CO_2_ concentration indoors. Results also led to the conclusion that the higher CO_2_ levels in the US setting were correlated to constant drowsiness reported in the home. This data highlights the importance of ventilation, even through the winter. The readings in the lengthy airplane journey confirmed that AMS’s readings and the airline’s advertised AQI were within range of each other. Figure 9 displays the time-series values of Carbon Monoxide [14].

**Fig 8.**
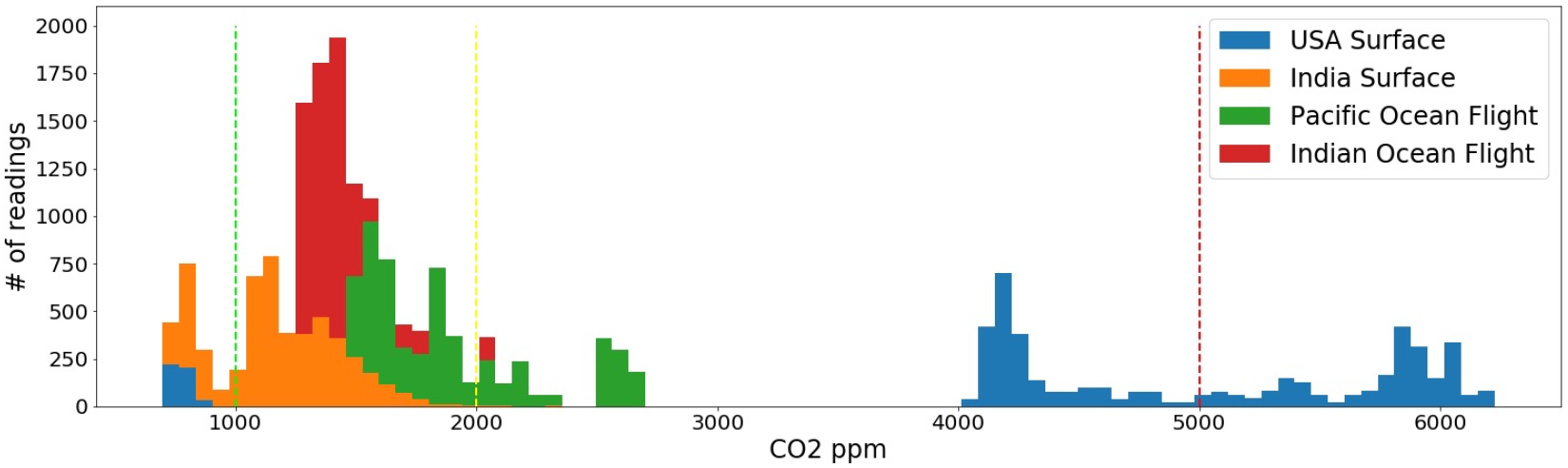
Carbon Dioxide (ppm): User atmosome in Cupertino, Hyderabad, a long airplane flight, and a shorter airplane flight. 0 (green vertical dash) to 1000 ppm (yellow vertical dash) is the healthy range, and anything higher than 5000 ppm is dangerous (red vertical dash). Comparison between short (red) and long (green) flights show the accumulation of cabin carbon dioxide which causes direct changes to blood bicarbonate levels.

**Fig 9.**
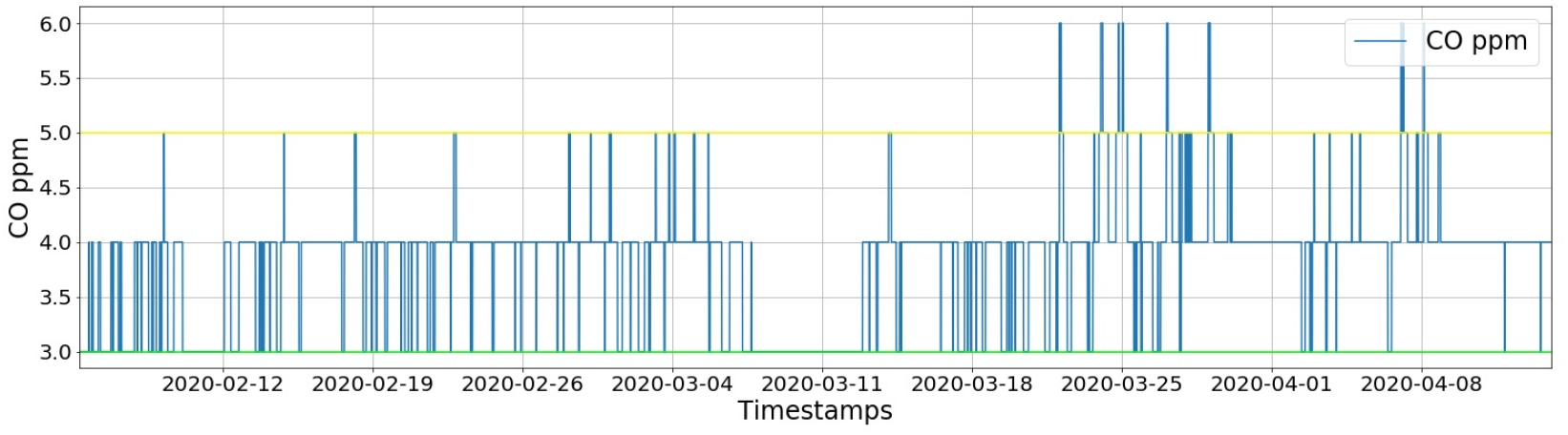
Carbon Monoxide (ppm): User home from February to April 2020 at home in the Sierra Mountains, El Dorado County, California. 0 to 3 ppm (green horizontal line) is the optimal range, and anything higher than 5 ppm (yellow horizontal line) is unhealthy.

### C. Particulate Matter 2.5µm (PM2.5)

Experiments measured particulate matter in 4 different conditions: daily residential life in the US, open traffic in India, daytime, and nighttime. The goal in the second condition was to see how the atmosome data, displayed in Figure 10, correlated with the throat infection that a participant sitting in an open 3-wheeler during the India traffic stretch developed after the journey. Analysis revealed that the particulate matter they inhaled, which exceeded the healthy recommendation, played a causal role in the sudden bout of throat and respiratory discomfort. The user’s cumulative PM2.5 exposure data is plotted in Figure 11. If a physician or user could access this kind of data regularly, they would gain insights on how their behavior and surroundings affect their bodies and take tangible steps towards staying healthy in traffic on vacation or other similar situations.

**Fig 10.**
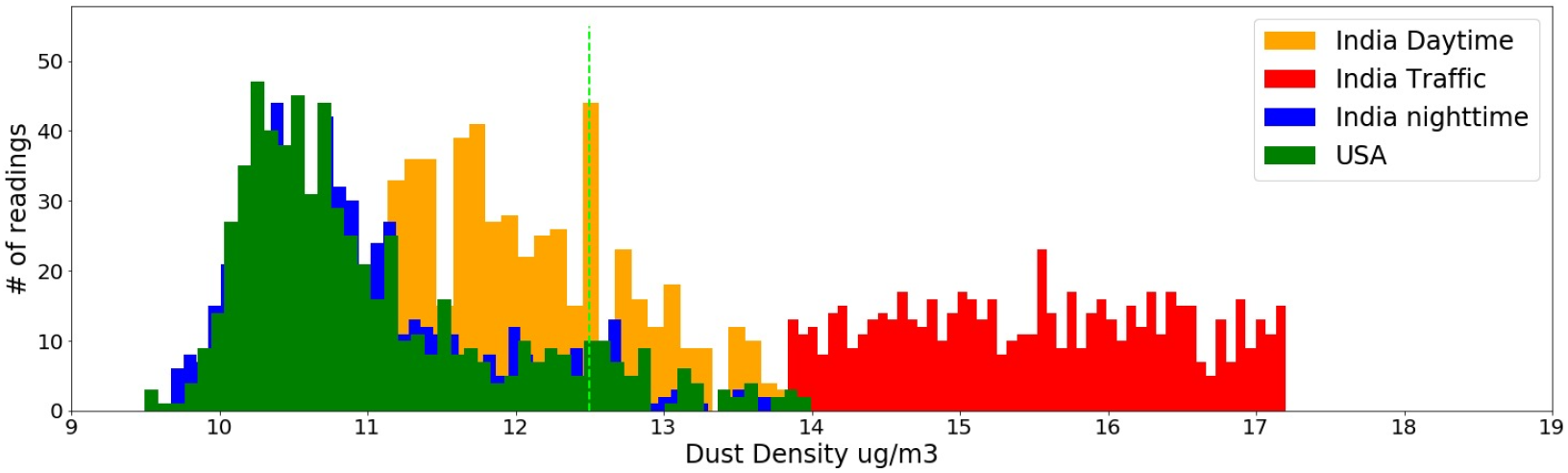
PM2.5 (in *µg/m*^3^): Personal atmosome in Hyderabad, India during the daytime, nighttime, and in traffic, and in Cupertino, California throughout the day in the home. 0 to 12.5 *µg/m*^3^ marks the healthy range (left of green vertical dash).

**Fig 11.**
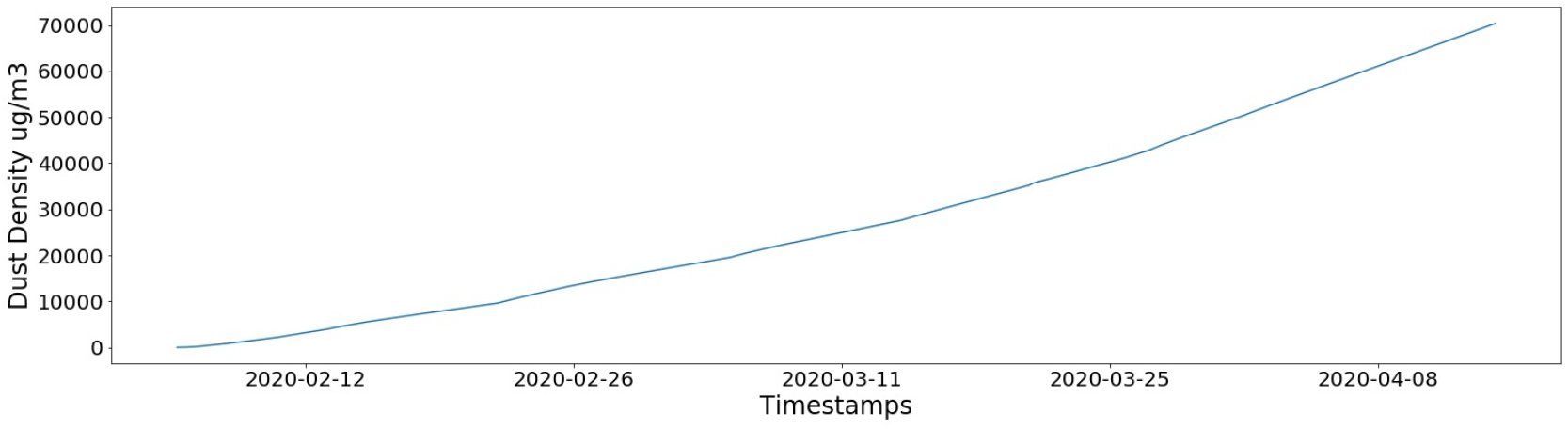
PM2.5 (in *µg/m*^3^) Cumulative Exposure: User home from February to April 2020 at home in the Sierra Mountains, El Dorado County, California.

### D. Volatile Organic Compounds (VOC) and Ozone

Figure 12 shows data collected by AMS for Total Volatile Organic Compounds (tVOC) and ground-level ozone. The time-series data, displayed in Figure 13 from the second user location, reflects unhealthy spikes in tVOC data on specific days. Evaluation by the user of potential indoor environmental stress factors on those days revealed that a running 3D printer in the user’s home caused these spikes. Further research confirmed that 3D printing is indeed a source of many VOCs [5]. Filament types varied from PLA, ABS, and TPU. This analysis established the need for extra ventilation when using a 3D printer.

**Fig 12.**
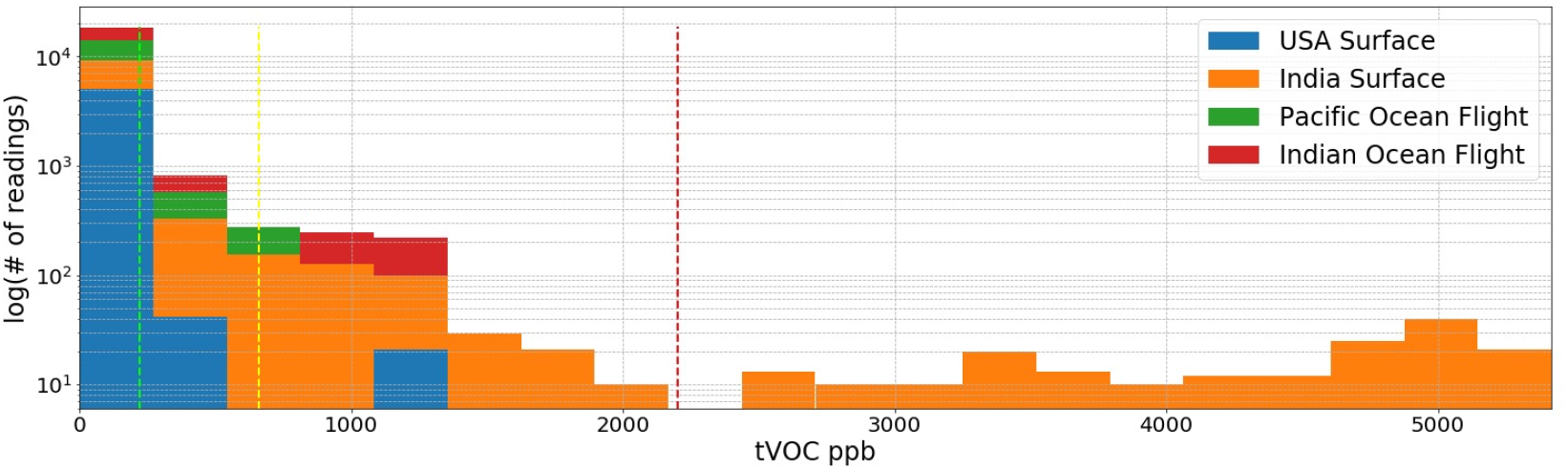
tVOC (ppb): User atmosome in 4 locations: Cupertino, California; Hyderabad, India; commercial airplanes over the Pacific; and commercial airplanes over the Indian Ocean. 0 to 200 ppb (left of green vertical dash) marks the healthy range, and anything above 2200 ppm (red vertical dash) is extremely unhealthy. We see that although commercial aircraft use filtration systems and are relatively high above surface air pollution, there is still a measurable effect on the cabin air quality based on ground air pollution (comparing green and red

**Fig 13.**
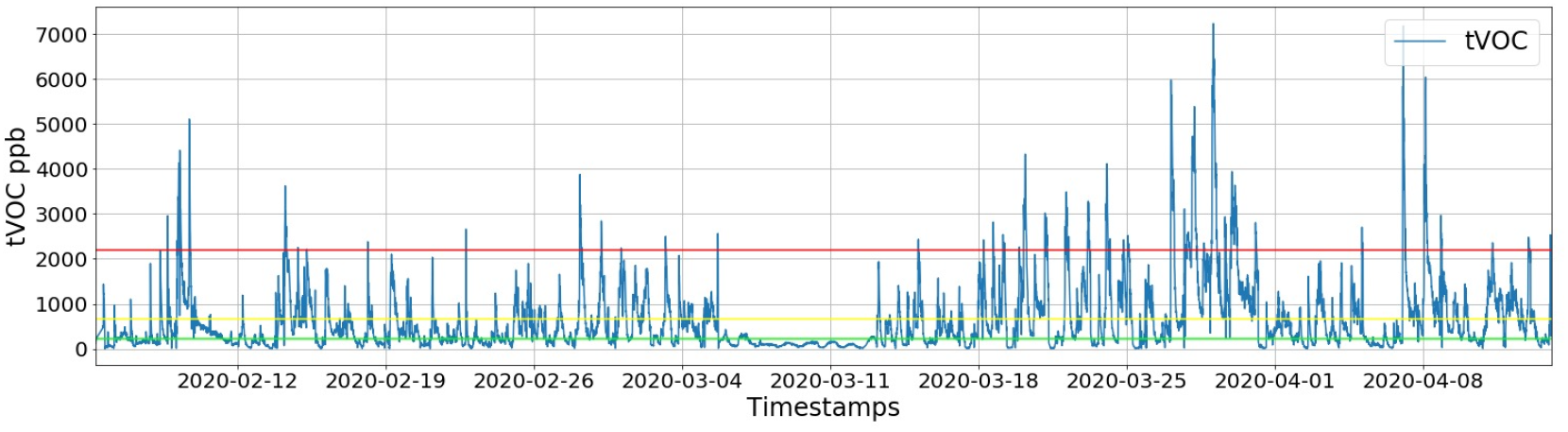
tVOC (ppb): User home from February to April 2020 at home in the Sierra Mountains, El Dorado County, California. 0 to 200 ppb (below green horizontal line) marks the healthy range, and anything above 2200 ppm is extremely unhealthy (above red horizontal line).

Figure 14 displays a strong correlation between tVOC and ground-level ozone. For the most part, spikes in tVOC result in increases in ground-level ozone, conforming with the chemical reactions that cause Nitrogen Oxides to react with tVOCs in the presence of sunlight to create ground-level ozone. This chemical process is demonstrated in Figure 15.

**Fig 14.**
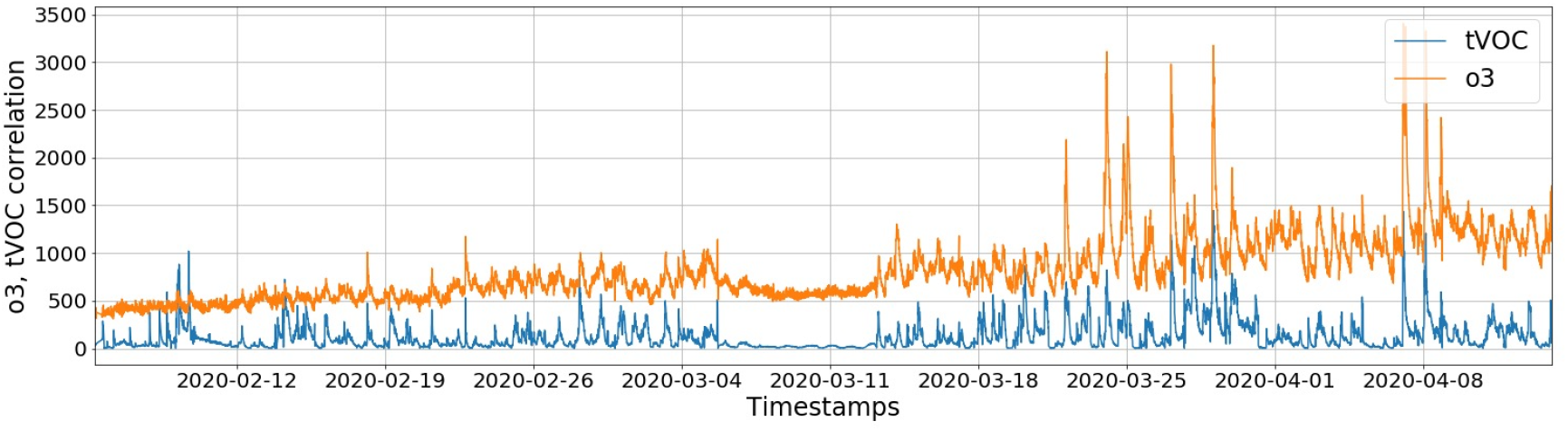
Ozone (ppb): User home ozone levels from February to April 2020 at home in the Sierra Mountains, El Dorado County, California.

**Fig 15.**
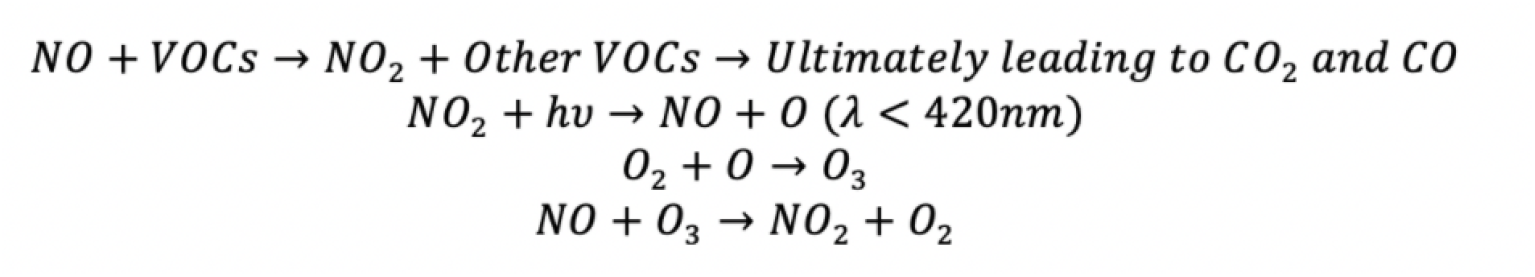
Ground Ozone Creation: Chemical reactions showing VOCs reacting with Nitrogen Oxides to create ground-level ozone.

Cumulative pollution data of tVOCs and ozone in the time-series user location shows how much tVOC and ozone pollution the user has been exposed to each day. Figures 16 and 17 show these plots. Acquisition of the 3D printer by the user shows how ozone and tVOC levels started to rise rapidly. Alerting the user of these changes can help with improving the personal atmosome.

**Fig 16.**
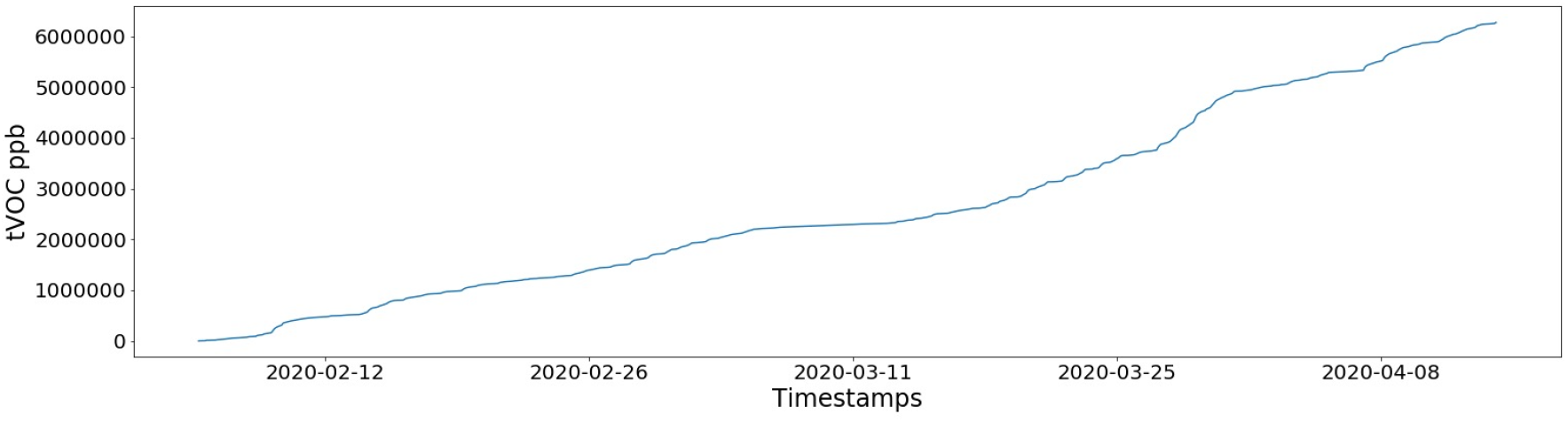
tVOC Cumulative Exposure: User home from February to April 2020 at home in the Sierra Mountains, El Dorado County, California.

**Fig 17.**
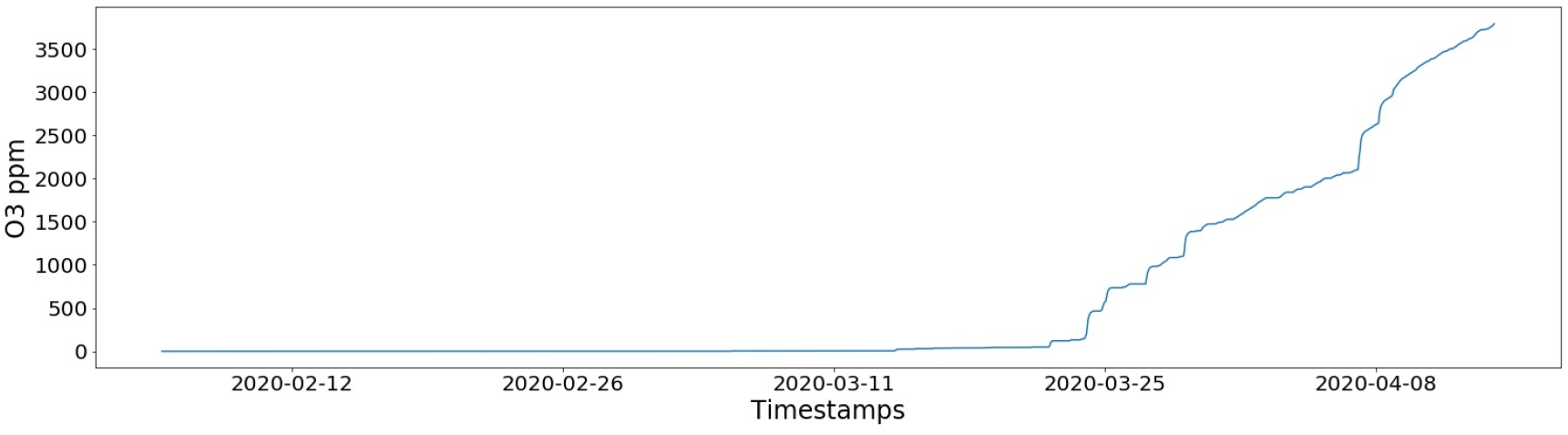
Ozone Cumulative Exposure: User home from February to April 2020 at home in the Sierra Mountains, El Dorado County, California.

## V. Discussion

The brief survey of currently available indoor air quality systems shows that AMS is one of the most comprehensive and low-cost IoT, indoor, real-time air monitoring systems. AMS also provides data accessible from anywhere in the world with internet connectivity. The data can be analyzed and compared across locations and time to identify patterns and provide recommendations to improve indoor environments. In this section, we will discuss how this data can directly reveal vital information about users’ health.

The atmosome, when combined with lifestyle and biological data collected from different wearable devices and smart phone applications presents a number of opportunities for personal health estimation and navigation [16], [15], [18].

We can perform N=1 experiments using the user’s lifestyle and atmosome data, and find the impact of atmospheric data streams on different aspects of user’s biology and behavior. The atmosome can also act as a confounding factor in many such relationships and thus should be further incorporated into the analysis.

### A. Temperature

Extreme changes in temperature increase mortality [25]. They also speed up the rate of some chemical reactions, including the rate at which ground-level ozone forms [9]. Our system is very effective in detecting such events, because of its capabilities to collect and post data to the cloud in real-time. Another characteristic that the system can give information on is brown adipose tissue metabolism. Colder temperatures correlate with activation of brown adipose tissue metabolism, which burns more calories [4]. The atmosome temperature sensor can collect data through the night to provide users and researchers with insight on their metabolism.

We characterize the direct impact of temperature reflected in user’s heart rate data stream during their sleep. Figure 18 shows the effect of ambient temperature during the night on a user’s heart rate while sleeping. This data was collected for one user over 424 nights of sleep using the iPhone mobile device with Sleep Cycle Application and Garmin Fenix 5 Watch. We clearly see increases in ambient temperature causes increases the user’s circulation flow (reflected by resting heart rate). Similar analyses can be performed to find the effect of other atmospheric streams on the user’s observed health and behavioral outcomes.

**Fig 18.**
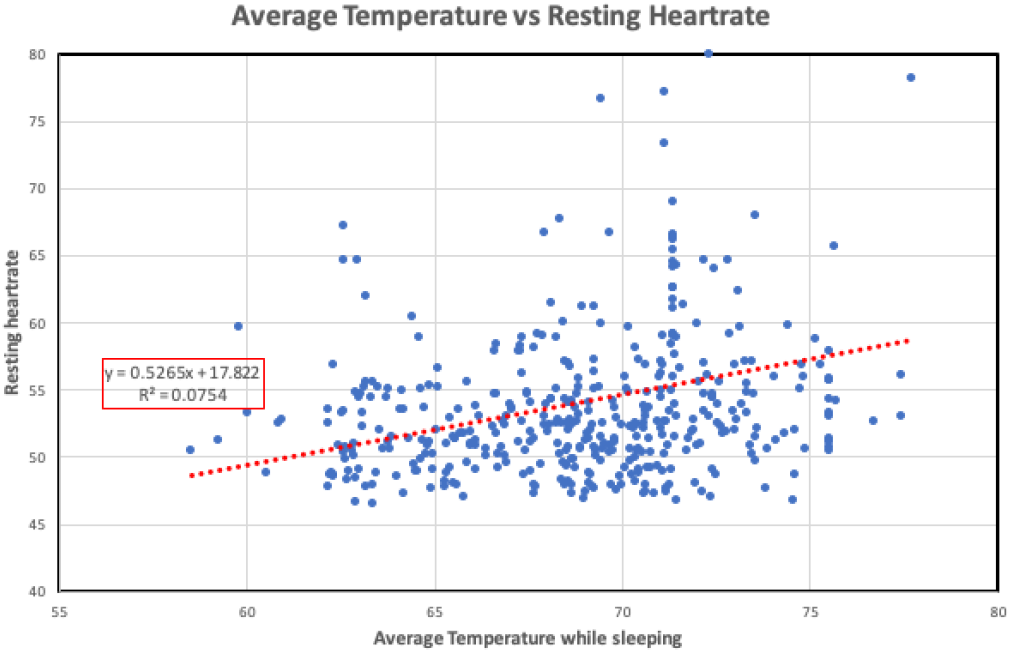
Personal Atmosome-Biology Modelling: This graph shows us how ambient temperature affects the user resting heart rate

### B. Humidity

Humidity affects skin dryness and corneal dryness as well as nasal passages and sinusitis [27]. Lower humidity levels can trigger the aforementioned conditions, while higher relative humidity levels promote the growth of mold, bacteria, and viruses. Inhaling mold particles leads to asthma attacks, respiratory infections, nose and eye irritations, and more [2]. Time series humidity data from AMS can provide users with valuable insight, such as whether to purchase a humidifier or take other actions to maintain an ideal health state.

Some of the health impacts of humidity can be seen in user input data streams. The measurable effects can include skin dryness, eye irritation, and can be recognized from images using image processing techniques such as color and line detection of redness and vessel structure on the sclera surface.

### C. Particulate Matter

Particulate matter refers to mixtures of microscopic solid and liquid particles suspended in air. There are two types of particulate matter that are most relevant to air pollution: PM10 and PM2.5. PM10 refers to particles that are between 2.5 and 10 microns; some examples of these include dust, pollen, and particles of mold. PM2.5 consists of fine particles that are 2.5 microns in diameter or less; fuel combustion, cigarette smoke, aerosols, and more can form them. Particulate matter is a health risk because it is small enough to be inhaled and deposits itself in airways of the human body. The smaller particles can even lodge themselves deep in the lungs or enter the bloodstream. Even short term exposure to PM10 has been associated with worsening respiratory diseases and can lead to emergency room visits. Long-term exposure (months to years) has been linked to premature death, especially in people with chronic conditions, and leads to reduced lung function in children [3]. WHO recommends a maximum exposure of 20 µg/m3 for PM10 and a maximum exposure of 10 µg/m3 for PM2.5 [31]. Measuring particulate matter in indoor air can lead to user implementable corrective actions. Users or researchers can view both event-specific and long-term PM levels, allowing them to assess the risks or benefits of specific lifestyle actions (i.e. being in an open vehicle in traffic) and their living environments at home (i.e. cooking practices and household chemical products).

We can observe the user’s response to particulate matter exposure using audio streams from a mobile phone or wearable device. The collected audio stream can be used to detect coughs [12], and allows us to monitor the changes in coughing, sneezing, wheezing and other audible symptoms in relation to changes in exposure to both PM2.5 and PM10 particles.

### D. Volatile Organic Compounds

A variety of household items, such as candles, cooking fumes, room fresheners, cosmetics, cleaning products, and paints, emit Volatile Organic Compounds (VOCs). VOCs are organic chemicals that are usually in gaseous form at room temperature, and they are photo-chemically active. Short-term exposure to VOCs can cause optic or respiratory irritation, headaches, memory lapses, and dizziness. Long-term exposure can also incite nausea, fatigue, organ damage, and cancer [20]. In our experimental results, we see that 3d printer use causes high spikes in tVOCs.

0–220ppb of tVOC is healthy, 221-660ppb is average, 661-2000ppb is dangerous, and greater than 2000 is very unhealthy [26]. AMS measures not just long-term general exposure, but also hyper-local short-term exposure through its collection of data every ten minutes.

### E. Carbon Dioxide

Carbon dioxide (CO_2_) classifies as a dominant air pollutant. Moderately high concentrations of CO_2_ in indoor air can lead to drowsiness, fatigue, and headaches, and increasing amounts can cause dizziness and nausea [19]. At extremely high concentrations, CO_2_ can even cause displacement asphyxia [23]. In our experimental data, we also see increases in CO_2_ during indoor exercise events. The simple process of improving ventilation can reduce high concentrations of CO_2_ in homes and workplaces and improve productivity and comfort.

Health effects of varying levels of CO_2_ are as follows: 351-450ppm is a healthy outdoor level, 451-700ppm is normal, 701-1000ppm is acceptable, 1001-2500ppm causes drowsiness and other symptoms of discomfort, 2501-5000ppm is detrimental to health, and anything higher than 5000ppm is extremely unhealthy [21].

### F. Ozone

While ozone in the stratosphere protects the Earth from the harmful rays of the sun, the ozone created at the ground level due to pollution is dangerous and can harm both humans and crops. Breathing ozone can trigger a variety of health problems, including chest pain, coughing, throat irritation, and airway inflammation. It can also worsen bronchitis, emphysema, and asthma, leading to the need for increased medical care [29]. In the United States in 2016, 90 percent of non-compliance to the national ambient air quality standards was due to ozone. Both short-term and long-term exposures to ozone, at concentrations below the current regulatory standards, are associated with increased mortality due to respiratory and cardiovascular diseases [33].

The guidelines for ozone exposure are the following: 0.05ppm is considered safe for up to 8 hours of exposure for someone doing heavy work outdoors. 0.08ppm is safe for up to 8 hours of exposure for someone doing moderate work outdoors. 0.1ppm is considered safe for up to 8 hours of exposure for someone doing light work outdoors. No more than 2 hours of exposure if safe for anyone for an ozone level of 0.2ppm or more [22].

## VI. Conclusions

In this paper, we present the concept of an atmospheric exposome — atmosome — and propose a low-cost approach to leverage multi-modal sensors in building a personal atmosome.

### Implications

The utility of this work is to create both the concept and system to quantify and measure the atmosphere that surrounds each individual in with continuosu real-time data streams and to give researchers access to this raw data though a cloud computing architecture. This system is useful for various audiences including user focused and research focused. For instance, it is important to detect smoke, CO, volatile compounds, gas leakage, and more in the houses of the elderly to avoid accidents that might result due to forgetfulness. AMS comes equipped to set up an email alert system to alert a caregiver immediately upon detecting any harmful levels of the indoor air and incite external action. It can also provide valuable feedback for medical professionals and researchers with both event-specific and time-series data.

### Limitations

AMS is limited in its capacity in various ways. This system at the current stage cannot yet give a validated prediction window of an adverse health event, such as cancer, that may happen. Although the Raspberry Pi has memory storage capabilities, we have not yet utilized these to allow the system to store data locally. Furthermore, the system is rather bulky at this stage, making it difficult to pocket or use as a constant tracker. By miniaturizing the system, we can incorporate it within mobile devices or wearable devices, thus giving a more unobtrusive experience for the user to collect more thorough and accurate atmosome data.

### Future Directions

Cybernetic and navigational health approaches enable individuals to be in control of their health throughout their lives and strives to maintain an ideal health state at all times [17], [16]. It consists of real-time, personalized data collection to pinpoint an individual’s current health state, followed by appropriate lifestyle recommendations to direct them towards their ideal state. Currently, the atmosome measurement system accomplishes the step in detecting events that impact the health state. In the future, we hope to develop a more robust health estimation and recommendation system that acts upon the atmosome data to advise individuals. This can help alter user behavior and lifestyle, especially regarding location and activities that affect atmosome health quality.

## Data Availability

Data is not publicly available.

## Notes

### Competing Interest Statement

The authors have declared no competing interest.

### Funding Statement

No external funding was received.

### Author Declarations

IRB approval was deemed not necessary for this research.

## References

[1] American Lung Association. Indoor Air Pollutants and Health — American Lung Association, 2020.

[2] American Lung Association. Mold and Dampness — American Lung Association, 2020.

[3] California Air Resources Board. Inhalable Particulate Matter and Health (PM2.5 and PM10) — California Air Resources Board, 2020.

[4] Andre’ C. Carpentier, Denis P. Blondin, Kirsi A. Virtanen, Denis Richard, François Haman, and Éric E. Turcotte. Brown adipose tissue energy metabolism in humans, 8 2018.

[5] Aika Y. Davis, Qian Zhang, Jenny P.S. Wong, Rodney J. Weber, and Marilyn S. Black. Characterization of volatile organic compound emissions from consumer level material extrusion 3D printers. Building and Environment, 160:106209, 8 2019.

[6] Dyson. Pure Hot + Cool, 2020.

[7] Paul J. Edelson, Phyllis E. Kozarsky, and Clive Brown. Air Travel - Chapter 8 - 2020 Yellow Book — Travelers’ Health — CDC, 2019.

[8] Environmental Protection Agency. Indoor Air Quality — EPA’s Report on the Environment (ROE) — US EPA, 2018.

[9] N. Fann, T. Brennan, P. Dolwick, J.L. Gamble, V. Ilacqua, L. Kolb, C.G Nolte, T.L. Spero, and L. Ziska. Ch. 3: Air Quality Impacts. The Impacts of Climate Change on Human Health in the United States: A Scientific Assessment. Technical report, U.S. Global Change Research Program, Washington, DC, 2016.

[10] Forbes. The 7 Best Air Purifiers For Your Home, 2020.

[11] Hannah Horvath. Best air purifiers of 2020, according to doctors and experts, 2020.

[12] Eric C. Larson, TienJui Lee, Sean Liu, Margaret Rosenfeld, and Shwetak N. Patel. Accurate and privacy preserving cough sensing using a low-cost microphone. In Proceedings of the 13th international conference on Ubiquitous computing - UbiComp ‘11, page 375, New York, New York, USA, 2011. ACM Press.

[13] Molekule. Molekule — Air Purifiers, 2020.

[14] My Home Comfort. Carbon Monoxide Levels & Risks - Home Comfort & Safety, 2020.

[15] Nitish Nag. Health State Estimation. PhD thesis, University of California, Irvine, 3 2020.

[16] Nitish Nag and Ramesh Jain. A Navigational Approach to Health: Actionable Guidance for Improved Quality of Life. Computer, 52(4):12–20, 12 2019.

[17] Nitish Nag, Vaibhav Pandey, Hyungik Oh, and Ramesh Jain. Cybernetic Health. Arxiv, 5 2017.

[18] Nitish Nag, Vaibhav Pandey, Preston J. Putzel, Hari Bhimaraju, Srikanth Krishnan, and Ramesh Jain. Cross-Modal Health State Estimation. In MM 2018 - Proceedings of the 2018 ACM Multimedia Conference, pages 1993–2002, New York, New York, USA, 10 2018. Association for Computing Machinery, Inc.

[19] National Institutes of Health. Carbon Dioxide: Your Environment, Your Health — National Library of Medicine, 2017.

[20] National Institutes of Health. Volatile Organic Compuounds (VOCs): Your Environment, Your Health — National Library of Medicine, 2017.

[21] Occupational Health and Safety. Carbon Dioxide Detection and Indoor Air Quality Control – Occupational Health & Safety, 2016.

[22] Occupational Safety and Health Administration. OSHA Occupational Chemical Database — Occupational Safety and Health Administration, 2020.

[23] Kris Permentier, Steven Vercammen, Sylvia Soetaert, and Christian Schellemans. Carbon dioxide poisoning: a literature review of an often forgotten cause of intoxication in the emergency department, 12 2017.

[24] Florina Pirlea and Wendy Ven-dee Huang. WDI - The global distribution of air pollution, 2019.

[25] Alexandra Schneider and Susanne Breitner. Temperature effects on health - current findings and future implications, 4 2016.

[26] Sensirion. Experts in Environmental Sensing Indoor Air Quality and Volatile Organic Compounds. Technical report, 2020.

[27] Shih Bin Su, Bour Wang, Chien Tai, Hsiu Fen Chang, and How Ran Guo. Higher prevalence of dry symptoms in skin, eyes, nose and throat among workers in clean rooms with moderate humidity. Journal of Occupational Health, 51(4):364–369, 2009.

[28] Dhruv Upadhyay, Vaibhav Pandey, Nitish Nag, and Ramesh Jain. N=1 Modelling of Lifestyle Impact on SleepPerformance. Arxiv, 6 2020.

[29] OAR US EPA. Ground-level Ozone Basics. Technical report, 2018.

[30] Jacinta C. Uzoigwe, Thavaleak Prum, Eric Bresnahan, and Mahdi Garelnabi. The emerging role of outdoor and indoor air pollution in cardiovascular disease, 8 2013.

[31] WHO World Health Organization. WHO Air quality guidelines for particulate matter, ozone, nitrogen dioxide and sulfur dioxide: Global update 2005. Technical report, 2005.

[32] WHO World Health Organization. WHO — Air pollution. WHO, 2017.

[33] Junfeng Jim Zhang, Yongjie Wei, and Zhangfu Fang. Ozone pollution: A major health hazard worldwide. Frontiers in Immunology, 10(OCT):2518, 10 2019.

